# Prefrontal hemodynamics during forward and backward walking, with and without a cognitive task, in people with MS: An fNIRS study

**DOI:** 10.1101/2022.10.18.22281209

**Authors:** Yana Kupchenko, Sapir Dreyer-Alster, Kim-Charline Broscheid, Alon Kalron

**Author notes:** **Correspondence:** A. Kalron, Department of Physical Therapy, School of Health Professions, Sackler Faculty of Medicine, and Sagol School of Neuroscience, Tel-Aviv University, Israel.

## Abstract

**Objective:** To investigate the prefrontal cortex (PFC) hemodynamics during forward and as well as backward walking, with and without a cognitive task, in people with MS (pwMS) and healthy controls.

**Methods:** The observational functional near-infrared spectroscopy (fNIRS) study comprised 18 pwMS and 17 healthy controls. Each subject completed four walking trials: Single task (ST) forward walking, dual task (DT) forward walking, ST backward walking, DT backward walking. PFC activity for all trials was recorded using functional near-infrared spectroscopy (fNIRS). The PFC was subdivided in the frontal eye field (FEF), frontopolar cortex (FPC) and the dorsolateral PFC (DLPFC).

**Results:** The relative oxygenated hemoglobin (HbO) concentration was higher during the DT forward walking in all PFC subareas compared with the ST forward walking for both groups. The relative HbO concentration was higher during ST backward walking compared with ST forward walking in pwMS (DLPFC, FEF) and the healthy controls (FEF, FPC), specifically during the initial part of the trial. There was no distinct difference in the relative HbO concentration between ST backward walking with DT backward walking in pwMS. In contrast, the relative HbO concentration in the FEF and the FPC increased in healthy controls during DT backward walking compared with ST backward walking, specifically during the second half of the trial.

**Conclusions:** ST backward walking and DT forward walking impact the hemodynamics at the PFC, although, the difference between pwMS and healthy adults requires further clarification. Future RCT’s are encouraged to examine the impact of an intervention program based on DT forward and backward walking on PFC activity in pwMS.

**Impact:** The present study demonstrated that backward walking increases activity in the PFC region. Similarly, when performing a cognitive task while walking forward. This information should be considered by PT’s for training, particularly pwMS.

## Introduction

Multiple sclerosis (MS), the most common chronic immune-mediated disorder affecting the central nervous system of young adults, is characterized by gait and debilitating cognitive impairments.^1-3^ MS-related gait difficulties contribute to a high risk of falling, as well as a reduced quality of life (QoL) and other negative real-world outcomes.^4,5^ Similarly, cognitive impairment, occurring in ∼50% of people with multiple sclerosis (pwMS),^6^ has been found associated with a host of negative consequences including depression, isolation, and a reduced QoL.^7-9^ Importantly, gait and cognitive impairments tend to co-occur in pwMS (i.e., cognitive-motor coupling).^10,11^

Previous studies have shown that pwMS possess an impaired ability to integrate cognitive and motor tasks (i.e., walking).^12^ Based on several systematic reviews, where pwMS were instructed to walk while performing a cognitive task (defined as a motor-cognitive dual task (DT)), their walking speed was reduced compared to standard walking (referred as the DT cost).^13-14^ Motor-cognitive DT involves an allocation of available attentional resources, requiring decision-making and executive functions. Consequently, the majority of studies investigating brain activity during motor-cognitive DT have focused on the prefrontal cortex (PFC), which plays a central role in cognitive control functions. Involvement of the PFC during motor-cognitive DT has been reported in healthy adults,^15^ stroke survivors,^16^ and people with Parkinson’s disease.^17^

Recently, a systematic review described neural-correlates of motor (walking)-cognitive DT in people with central neurological disorders;^18^ six of these studies investigated pwMS,^19-24^ only one provided information on the PFC.^24^ Hernandez et al reported that the increase in PFC oxygenation concentration during motor (walking)-cognitive DT (compared to single task (ST) walking) was greater in pwMS compared with the healthy controls.^24^ Worth noting, only two (out of six) studies^21,24^ assessed brain activity by a functional near infra-red spectroscopy device (fNIRS). This device is a non-invasive brain imaging tool to measure blood oxygenation change and is a common system used to monitor brain activity during motor-cognitive DT. fNIRS has better temporal resolution, is easier to use, and more cost effective to implement than an fMRI, although, spatial resolution is more limited. Additionally, the fNIRS has better spatial resolution and is more robust against motion artifacts than an electroencephalogram (EEG), although temporal resolution is more limited.^25^

In the present study, we explored further the role of the PFC during motor-cognitive DT by supplementing backward walking to forward walking. Backward walking is crucial when performing activities of daily living such as backing up to a chair or moving away from the refrigerator door when closing it. Previous studies have recognized that backward walking can serve as a sensitive clinical measure of mobility in the elderly^26^ including those with dementia,^27^ and pwMS.^27,28^ Backward walking requires increased cognitive demands and postural control^29^ vis a vis forward walking. In pwMS, deficits in balance increase during backward walking and significantly correlate with severity regarding clinical measures of mobility and disability.^30^ Furthermore, pwMS, when administered with a cognitive DT, display more prominent deficits during backward walking than during normal walking.^31,32^

A question of interest is whether ST forward walking and ST backward walking require different neural activity at the PFC. Moreover, the effect of a concurrent cognitive DT during backward walking on the neural activity in the PFC, is unknown. Data on this issue might be beneficial for pwMS since backward walking maybe successfully applied in rehabilitation therapy.^33^

Therefore, the aim of our study was to investigate the PFC hemodynamics during forward and backward walking during ST and DT in pwMS versus healthy controls. Based on the literature, our hypotheses were that PFC hemodynamics would differ between ST and DT, in both forward and backward conditions, in both groups, however, to a larger extent in pwMS. Backward walking would result in differences in the PFC hemodynamics compared with forward walking during ST and DT conditions in both pwMS and healthy controls.

## Materials and methods

### Study design and participants

This observational fNIRS case-control study was conducted at the Multiple Sclerosis Center, Sheba Medical Center, Tel-Hashomer, Israel and comprised a convenience sample of 18 pwMS (36.1 ± 11.7 years, 66.6% female) and 17 healthy controls (37.5 ± 13.8 years, 76.5% female). Due to lack of comparable studies, no sample size calculation was performed. However, our sample size was similar to other fNIRS gait studies researching pwMS.^20,21,24^ pwMS were enrolled in the study according to the following inclusion criteria: (1) a neurologist-confirmed diagnosis of definite MS according to the revised McDonald criteria;^34^ (2) ability to walk without a cane or canes or walker or rollator. Exclusion criteria included: (1) corticosteroid treatment within 90 days prior to examination; (2) other significant neurological or psychiatric illnesses; (3) diagnosed with dysregulation of the autonomic nervous system; (4) blood pressure irregularities (e.g. prehypertension, hypertension, orthostatic hypotension) (self-report); (5) alcohol or drug abuse, and (6) orthopedic disorders that could negatively affect walking. The objectives and requirements of our study were explained to all the participants who prior to their participation provided informed consent. Approval was obtained from the Sheba Medical Center Independent Ethics Committee before commencement of the study (Ethics Ref: 5596-08/141210).

### Experimental procedure

The experiment was conducted at a single session in Sheba MSC. Following the acquisition of informed consent, the participants completed the Symbol Digit Modalities Test (SDMT), a measure of information processing,^35^ and the 12-item Multiple Sclerosis Walking Scale (MSWS-12), a measure of patient perception of the impact of MS on walking ability.^36^ The experiment was continued in a separate room embedded with a 25 m walking path. Each subject completed a sequence of four consecutive walking trials with a 1 min rest break between trials. All walking trials were performed with the subject wearing an fNIRS device (a cap and a 2 kg backpack (NIRSport2, NIRx Medical Technologies LLC, New York, USA)). Each walking trial was repeated 3 times (30 s each trial, with a rest interval of 40 s). The trials consisted of the following conditions:

1. ST forward walking: All participants wore their everyday footwear and were instructed to walk across the room at their normal walking speed. At the end of the walking path, the participant was instructed to turn around and continue until the end of the 30 s trial, announced aloud by the tester.
2. DT forward walking: The procedure was identical to the ST forward walking except that subjects were instructed to perform the Serial-7 Backward Test^37^ while walking. They were asked to count backwards aloud in increments of seven starting from a randomly chosen 3-digit number between 300 and 400.
3. ST backward walking: The condition was also identical to normal walking except for walking backward at a self-selected pace while looking straight ahead. The tester walked alongside the subject for safety, informing the patient when the end of the path was reached.
4. DT backward walking: The subjects were instructed to perform the Serial-7 Backward Test while walking backward. During the rest intervals, the subject was instructed to stand in place with a steady walker to lean on. Furthermore, in order to minimize the influence of mind wandering on PFC brain activity, the subject was instructed to count quietly upwards by one during the resting period. The experiment was coded by the Python PsychoPy® software (Open Science Tools Ltd).^38^ PsychoPy® (https://psychopy.org/index.html) is an application for creating experiments in behavioral sciences with precise spatial control and timing of stimuli. The coded experiment, including triggers for the walking trials, was synced with the recording signal of the fNIRS system.

### Equipment and Outcome Measures

For this study, a portable fNIRS system (NIRSport, NIRx Medical Technologies, NY, USA) was attached to a standardized cap (EasyCap GmBH, Herrsching, Germany), 58 cm in circumference, and equipped with eight sources and eight detectors with eight short separation channels according to the International 10–20 system for EEG to cover the PFC. The average source-detector separation distance was 30–40 mm. The arrangement of the optodes was performed with the fNIRS Optodes’ Location Decider (fOLD) toolbox.^39^ Additional information as to the sensitivity of the channels according to the fOLD toolbox is provided in the supplementary material. The cap was placed in the middle of the scalp between nasion and inion and between the left preauricular and the right preauricular point (reference point Cz). The applied fNIRS system operates at two different wavelengths (760/850 nm) and at a fixed sampling frequency of 7.81 Hz. The subareas captured are the dorsolateral PFC (Brodmann area 9) (DLPFC), frontopolar PFC (Brodman area 10) (FPC) and the frontal eye fields (Brodman area 8) (FEF). A fair to excellent inter-session reliability of the fNIRS-derived parameters in the subareas of the PFC measured while walking has been proven in healthy controls.^40^ In order to control physiological fNIRS signal confounders we used eight short-separation channels (∼8mm) which is sensitive to blood perfusion and oxygenation changes in the extracerebral tissue layer. Additionally the Polar H10 sensor chest strap device was used to determine heart rate as well as heart rate variability.

### Functional Near-Infrared Spectroscopy Data Processing

To process and convert the fNIRS data, Homer3 (version 1.32.4) was used.^41^ Non-existing values were replaced by spline interpolation (function hmrR PreprocessIntensity NAN). Channels with a too weak or too strong signal as well as a too high standard deviation were excluded (function: hmrR PruneChannels: data range = 1 × 10^−2^ to 1 × 10^7^; signal-to-noise threshold = 2; source detector separation range: 0.0–45.0 mm). The preprocessed raw data were then converted to optical density data (function: hmR Intensity2OD).^41^ Using the spline interpolation and a digital Savitzky-Golay filter motion, artifacts were removed (function: hmR MotionCorrectSplineSG: p = 0.99; frame size = 15 s).^42^ The 3^rd^ order Butterworth bandpass filter was applied to diminish physiological artifacts (function: hmrR BandpassFilt: Bandpass Filter OpticalDensity).^43^ Therefore, the high-pass filter was set at 0.01 Hz to minimize the proportion of oscillations associated with the vascular endothelial function.^43^ The low-pass filter was set at 0.09 Hz to primarily filter out the Mayer waves.^44^ Subsequently, the optical density data were converted to concentration data by the Beer-Lambert law adapting the differential path length factor to the age of each participant.^45^ Finally, the individual hemodynamic response function (HRF) was calculated with the ordinary least squared deconvolution method by utilizing a general linear model approach (function: hmrR GLM).^46^

The HRF was based on a consecutive sequence of Gaussian functions (width of the Gaussian 0.5 and temporal spacing between the consecutive Gaussian 0.5). The short separation regression was performed with the nearest short separation channel. The 3^rd^ order polynomial drift baseline correction was applied. Subsequently, the data were processed by the MATLAB program (version R2020b, The MathWorks, Natick, Massachusetts, USA). Initially, the early phase of task onset (5 s) was eliminated for each subject to avoid transient effects of movement initiation on the hemodynamic response.^40^ Secondly, the last 5 s were eliminated to minimize the impact of the expected ending of the walking trial. Accordingly, data recorded during the time interval of 5–25 s from each walking trial, were analyzed. The relative oxygenated hemoglobin (HbO) and deoxygenated hemoglobin (HbR) concentration data of each channel during this time interval were then averaged for each subject. Finally, the channels were merged into the subareas of the PFC as described above.

### Gait measures

Gait was evaluated via three small, lightweight axial wearable accelerometers (APDM, Oregon, USA) positioned on the dorsum of both feet and at the level of the lumbosacral junction, attached with elastic straps. The sensors and the respective APDM’s Mobility LabTM software analyzed the spatio-temporal parameters of gait. This system is both accurate and repeatable for measuring spatio-temporal gait parameters.^47,48^ Since the focus of the present study was on PFC activity during walking, only the walking speed was analyzed.

### Statistical analysis

Descriptive statistics determined the demographics, walking speed, clinical characteristics, and hemodynamic PFC measures of the study sample. Normal distribution was verified using the Kolmogorov-Smirnov test. Additionally, the relative HbO and HbR concentration data were examined with boxplots for each subarea of the PFC (DLPFC, FPC and FEF). In the event of outlier identification, the data were removed. Study groups were compared for age, walking speed and cognitive status (represented by the SDMT) by the t and chi-square test for gender. A general linear model performed repeated measures analysis of variance on HbO and HbR concentration data.

All analyses were performed using the SPSS software (version 27.0 for Windows; SPSS Inc., Chicago, Illinois, USA). All reported p-values were two-tailed. The level of significance was set at p < 0.05. The figures illustrating the relative HbO concentration change in the PFC subareas during the four walking trials for each group were created using the R software program (R studio version 1.4.1717). The Ggplot2 library was utilized to create the plots by implementing the geom_smooth function to produce a curve estimating the conditional mean function. The generalized additive model method was applied.

## Results

The clinical characteristics of the 35 participants are summarized in Table 1. No significant differences were observed between pwMS and healthy controls in terms of age and gender. The median Expanded Disability Status Scale score of the pwMS was 3.0 indicating mild disability; the mean disease duration from diagnoses was 7.5 (S.D.=4.9) years. All pwMS had undergone a relapsing-remitting clinical disease course. Significant differences between groups were observed for walking speeds in all four conditions. pwMS walking speed was ∼14% and ∼28% slower during ST forward and ST backward walking, respectively, compared with healthy controls. Moreover, walking speed was slower by 51% and 41% during ST backward walking compared with ST forward in pwMS and healthy controls, respectively. Furthermore, the addition of a cognitive DT reduced walking speed in both forward and backward walking conditions. Walking speed during DT forward was slower than ST forward by 11% and 9% in pwMS and healthy controls, respectively. Walking speed during DT backward was slower than ST backward by 30% and 18% in pwMS and healthy controls, respectively.

**Table 1.** Demographical and clinical data of the study sample

|  | MS<br>(n=18) | Healthy controls<br>(n=17) | *p*-value |
| --- | --- | --- | --- |
| Age, y | 36.1 (11.7) | 37.5 (13.8) | 0.876 |
| Gender, f/m | 12/6 | 13/4 | 0.453 |
| Disease duration, y | 7.5 (4.9) | --- | --- |
| Median EDSS, score [range] | 3.0 [1.0-5.0] | --- | --- |
| SDMT, score | 48.2 (12.9) | 55.9 (11.2) | 0.146 |
| MSWS-12, score | 24.7 (6.3) | --- | --- |
| *Walking speed, m/s* |  |  |  |
| ST forward | 1.22 (0.27) | 1.41 (0.15) | **0.032** |
| DT forward | 1.08 (0.42) | 1.28 (0.25) | **<0.001** |
| ST backward | 0.60 (0.23) | 0.83 (0.14) | **<0.001** |
| DT backward | 0.42 (0.19) | 0.68 (0.18) | **<0.001** |

Scores are presented as mean (S.D.)
\*MS, multiple sclerosis; y, years; f, females; m, males; EDSS, Expanded Disability Status Scale; SDMT, Symbol Digit Modality Test; MSWS-12, MS walking scale 12 items; ST, walking single-task; DT, walking-cognitive dual-task;
Bold: $p$ -value < 0.05;

The relative HbO and HbR concentration in the PFC subareas according to the four walking conditions and groups are presented in Table 2. No differences were found in the hemodynamic measures in the PFC subareas between the four walking conditions in pwMS. Significant differences were observed between ST forward and DT forward walking for HbO concentration (−0.081 (S.D.=0.199) μmol/l vs. 0.101 (S.D.=0.221) μmol/l; p < 0.001) and HbR concentration (0.047 (S.D.=0.081) μmol/l vs. 0.001 (S.D.=0.053) μmol/l; p = 0.038) in the FEF in the healthy controls. Also, a significant difference was found in the relative HbO between ST forward with DT forward walking (−0.281 (S.D.=0.291) μmol/l vs. 0.064 (S.D.=0.439) μmol/l; p < 0.001) in the FPC in the healthy controls. In terms of the total group (pwMS and healthy controls), a significant difference was found between ST forward with ST backward walking (p = 0.039) and DT backward walking (p = 0.029) in the relative HbO concentration in the FPC. Moreover, a significant difference was found between ST forward and DT forward walking (p = 0.001) in the relative HbO concentration in the FEF. Non-significant scores were observed for group times walking condition in the relative HbO and HbR concentration measures.

**Table 2.** Mean (S.D.) of the relative oxy and deoxyhemoglobin concentrations in the subareas of the prefrontal cortex during all four walking conditions in the study groups.

| Group | Healthy controls (n=17) |  |  |  |  | MS (n=18) |  |  |  |  | Condition |
| --- | --- | --- | --- | --- | --- | --- | --- | --- | --- | --- | --- |
| Condition | ST forward | DT forward | ST backward | DT backward | *p*-value | ST forward | DT forward | ST backward | DT backward | *p*-value | *p*-value |
| *DLPFC, Dorsolateral prefrontal cortex (Brodmann area 9)* |  |  |  |  |  |  |  |  |  |  |  |
| HbO<br>[μmol/l] | -0.051<br>(0.251) | 0.069<br>(0.388) | 0.024<br>(0.293) | 0.056<br>(0.264) | 0.399 | -0.149<br>(0.246) | -0.062<br>(0.221) | -0.015<br>(0.314) | -0.011<br>(0.316) | 0.375 | 0.091 |
| HbR<br>[μmol/l] | 0.020<br>(0.071) | 0.015<br>(0.087) | 0.012<br>(0.084) | -0.063<br>(0.127) | 0.423 | -0.030<br>(0.067) | 0.002<br>(0.042) | 0.028<br>(0.080) | 0.002<br>(0.096) | 0.457 | 0.723 |
| *FPC, Frontopolar prefrontal cortex (Brodmann area 10)* |  |  |  |  |  |  |  |  |  |  |  |
| HbO<br>[μmol/l] | -0.281<sup>a</sup><br>(0.291) | 0.064<sup>b,c</sup><br>(0.439) | 0.097<sup>a,b,c</sup><br>(0.360) | 0.080<sup>b,c</sup><br>(0.291) | **<0.001** | -0.079<br>(0.319) | -0.056<br>(0.328) | -0.101<br>(0.334) | -0.094<br>(0.293) | 0.913 | **0.011<sup>x</sup>** |
| HbR<br>[μmol/l] | 0.048<br>(0.134) | -0.016<br>(0.153) | 0.008<br>(0.097) | 0.032<br>(0.095) | 0.223 | -0.009<br>(0.109) | -0.035<br>(0.090) | -0.028<br>(0.092) | -0.025<br>(0.072) | 0.846 | 0.195 |
| *FEF, Frontal eye fields (Brodmann area 8)* |  |  |  |  |  |  |  |  |  |  |  |
| HbO<br>[μmol/l] | -0.081<sup>a</sup><br>(0.199) | 0.101<sup>b</sup><br>(0.221) | -0.003<sup>a,b</sup><br>(0.216) | 0.072<sup>b</sup><br>(0.247) | **<0.001** | -0.091<br>(0.312) | 0.073<br>(0.371) | 0.032<br>(0.320) | 0.011<br>(0.283) | 0.286 | **0.002<sup>y</sup>** |
| HbR<br>[μmol/l] | 0.047<sup>a</sup><br>(0.081) | 0.001<sup>b</sup><br>(0.053) | 0.014<sup>a,b</sup><br>(0.068) | 0.037<sup>a,b</sup><br>(0.131) | **0.038** | -0.018<br>(0.065) | -0.007<br>(0.071) | -0.010<br>(0.086) | 0.024<br>(0.096) | 0.351 | 0.693 |

Bold, *p*-value<0.05,
a,b,c indicate significant differences between walking conditions.
x; significant difference between NW and BW/BWDT; y; significant difference between ST forward and DT forward

The time course of the mean relative HbO concentration over the walking trials in each group is displayed in Figures 1-3. The relative HbO concentration was higher during ST backward walking compared with ST forward walking in pwMS (DLPFC, FEF) and the healthy controls (FEF, FPC), specifically during the initial part of the trial (0–10 s) (Figure 1a-c). The relative HbO concentration was higher during the DT forward walking in all PFC subareas compared with the ST forward walking for both groups (Figures 2a-c). There was also a larger reduction in the relative HbO concentration in the DLPFC and FEF during the second half of the trial (15–30 s) in pwMS compared with healthy controls. As for backward walking (Figures 3a-c), there was no distinct difference in the relative HbO concentration between ST backward walking with DT backward walking in pwMS. In contrast, the relative HbO concentration in the FEF and the FPC increased in the healthy controls during DT compared with ST backward walking, specifically during the second half of the trial (15–30 s).

**Fig. 1a.**
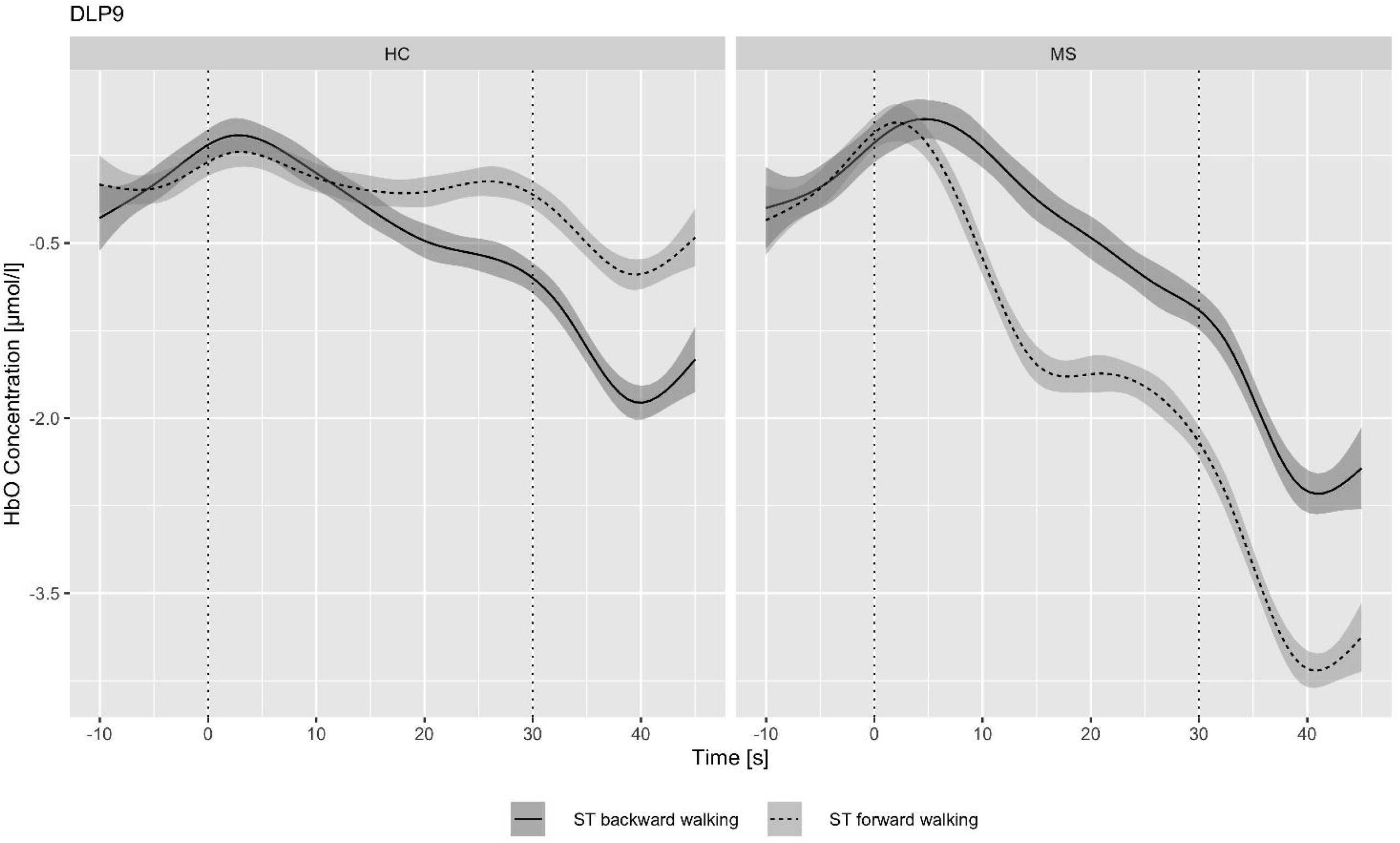
HbO concentration in the DLPFC during ST forward and ST backward walking in pwMS (MS) and healthy controls (HC)

**Fig. 1b.**
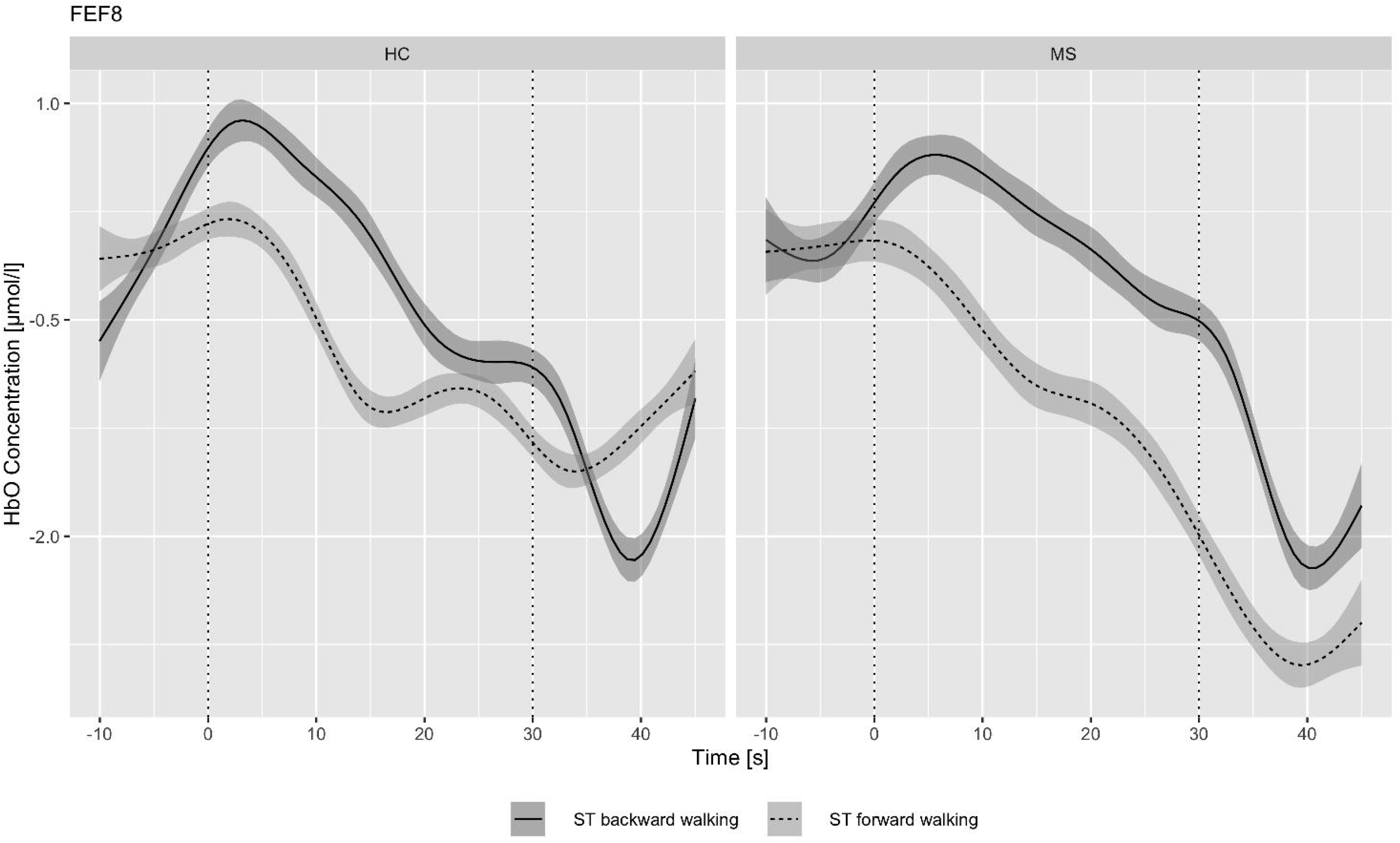
HbO concentration in the FEF during ST forward and ST backward walking in pwMS (MS) and healthy controls (HC)

**Fig. 1c.**
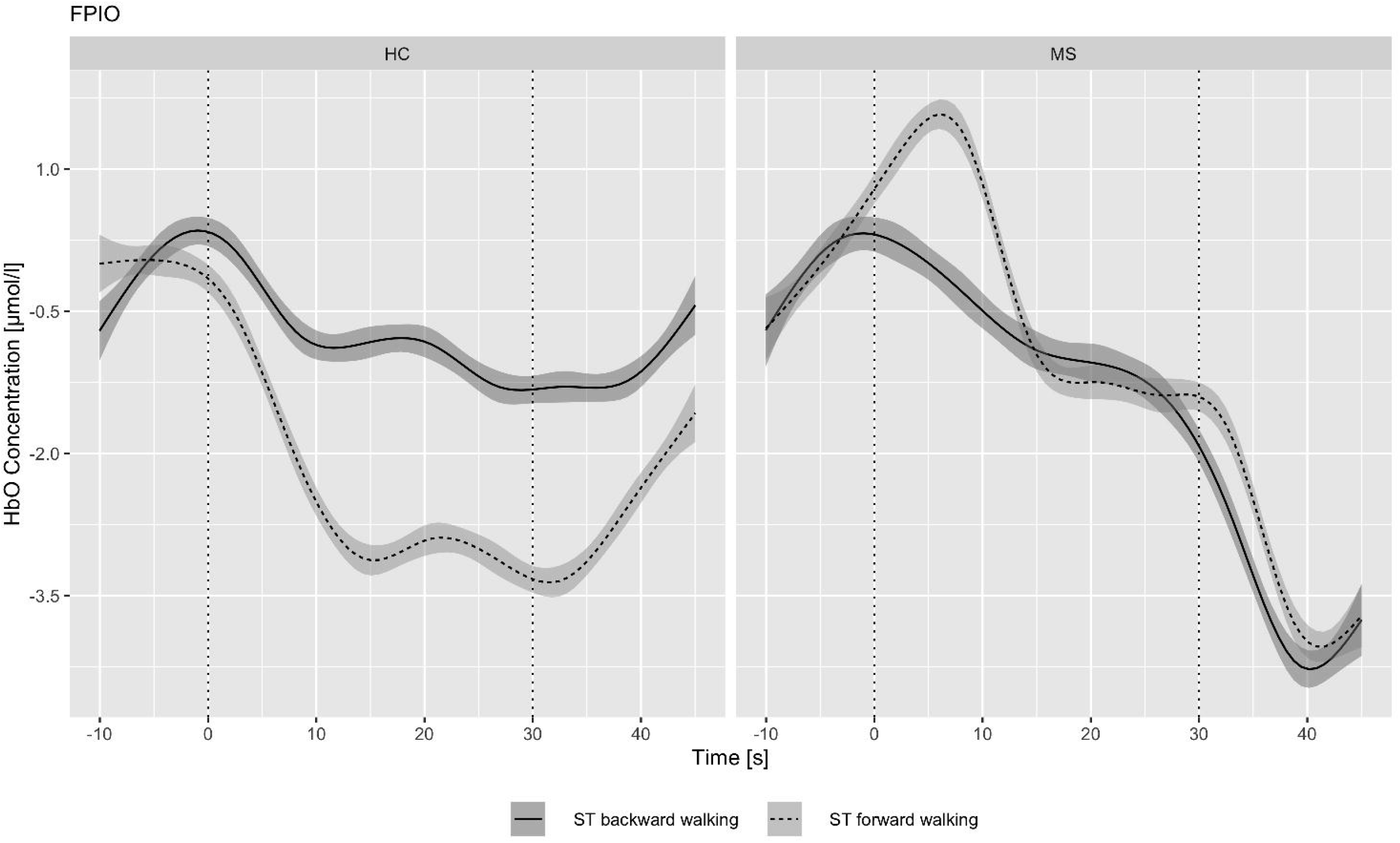
HbO concentration in the FPC region during ST forward and ST backward walking in pwMS (MS) and healthy controls (HC)

**Fig. 2a.**
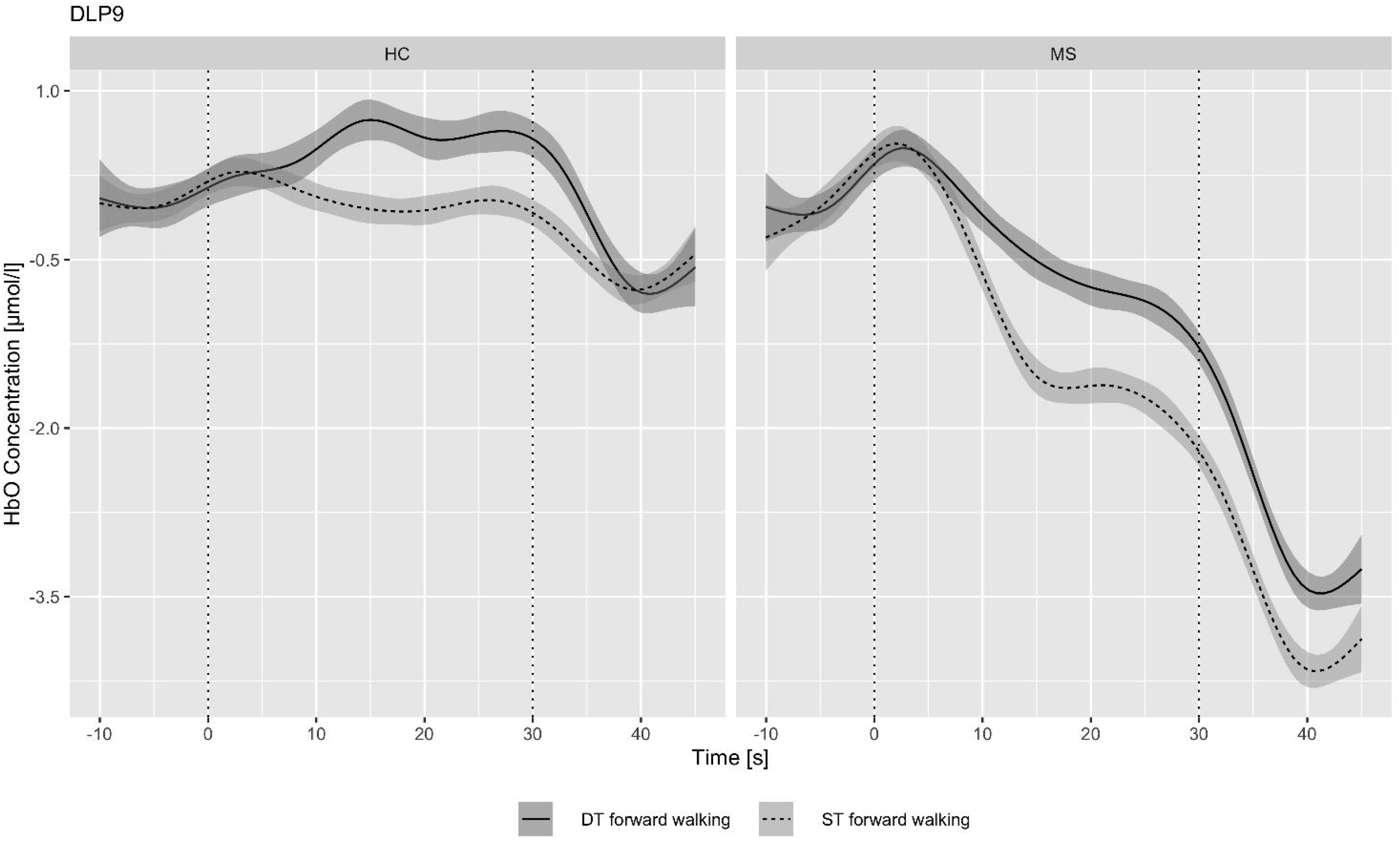
HbO concentration in the DLPFC during ST forward and DT forward walking in pwMS (MS) and healthy controls (HC)

**Fig. 2b.**
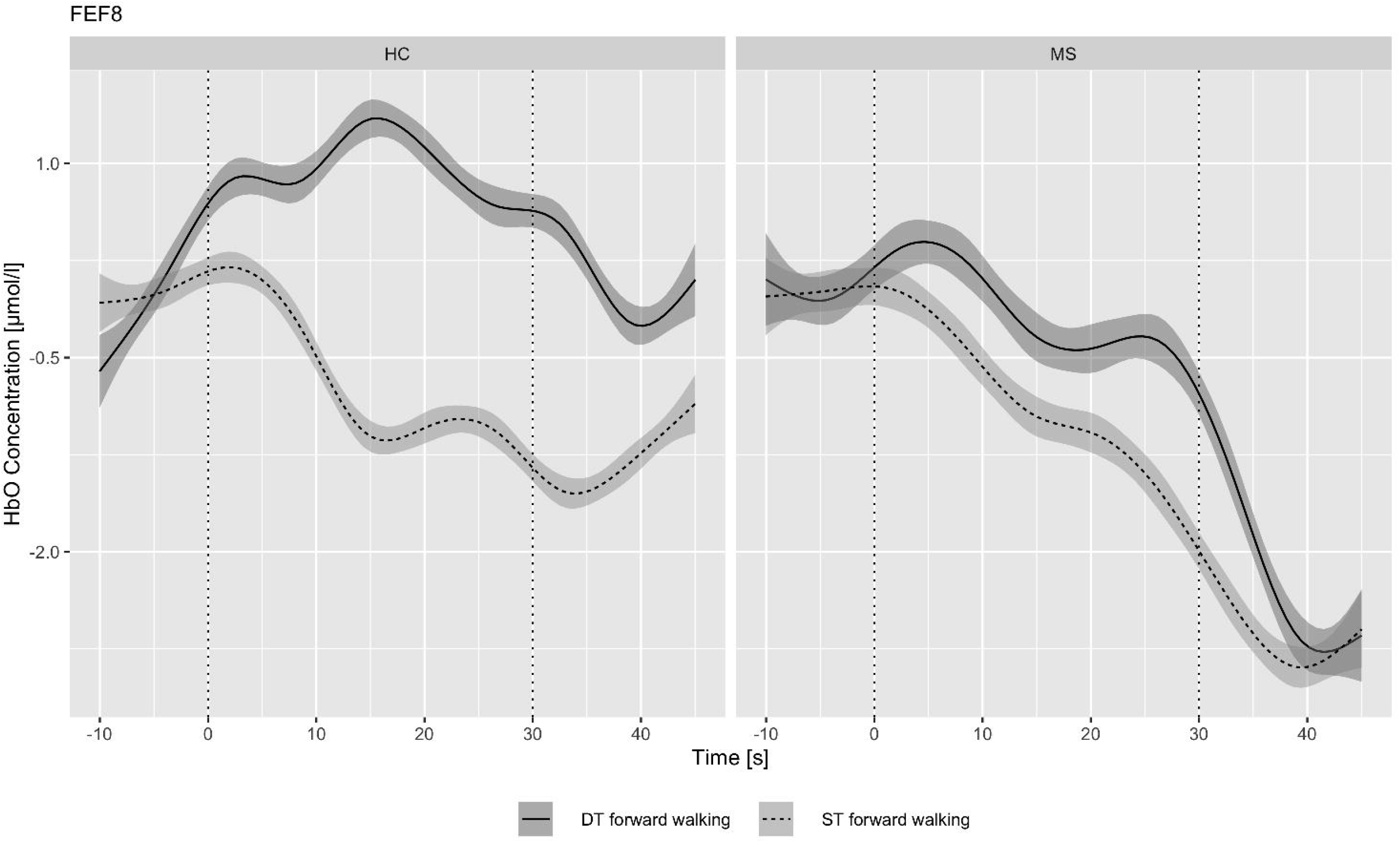
HbO concentration in the FEF during ST forward and DT forward walking in pwMS (MS) and healthy controls (HC)

**Fig. 2c.**
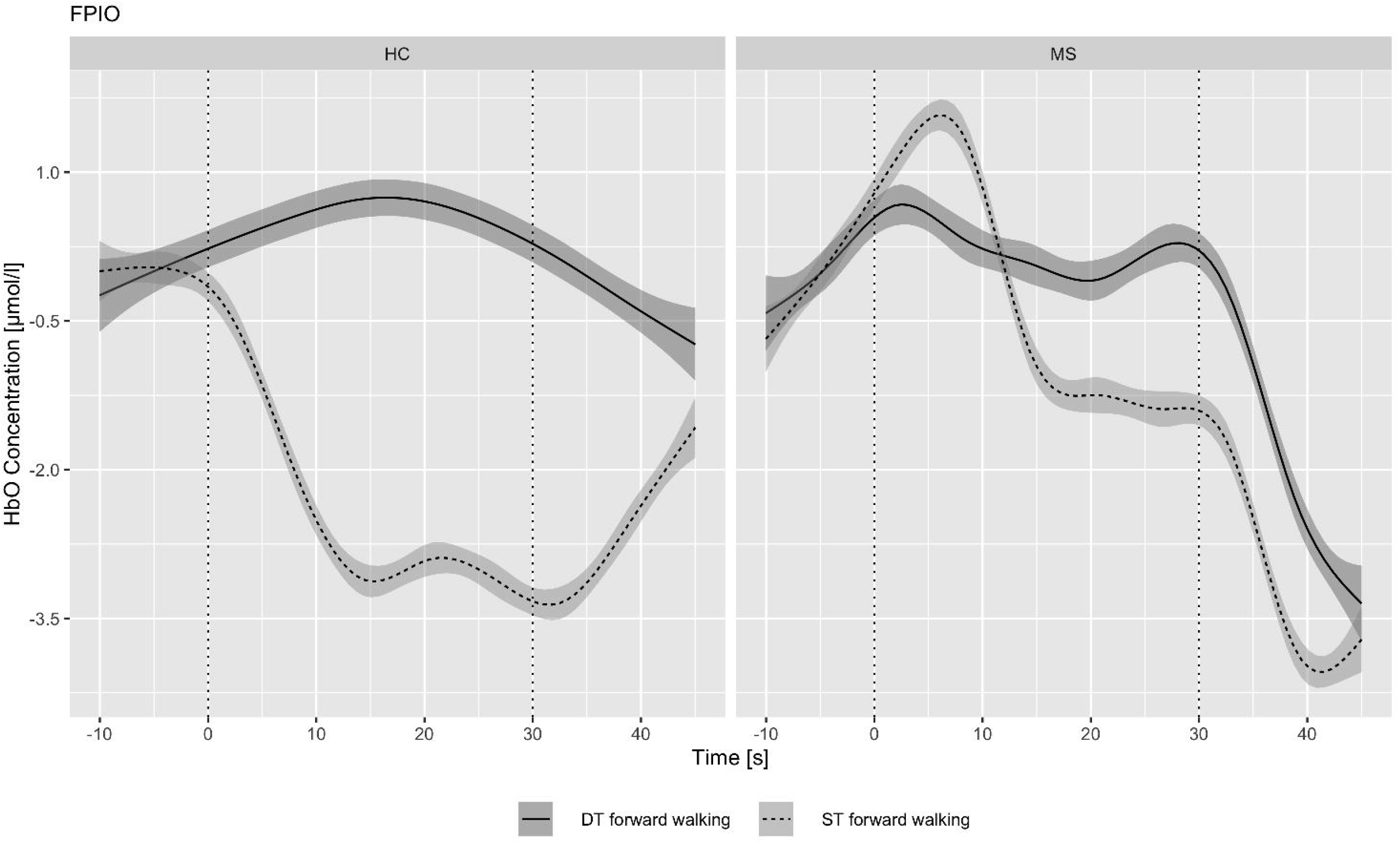
HbO concentration in the FPC during ST forward and DT forward in pwMS (MS) and healthy controls (HC)

**Fig. 3a.**
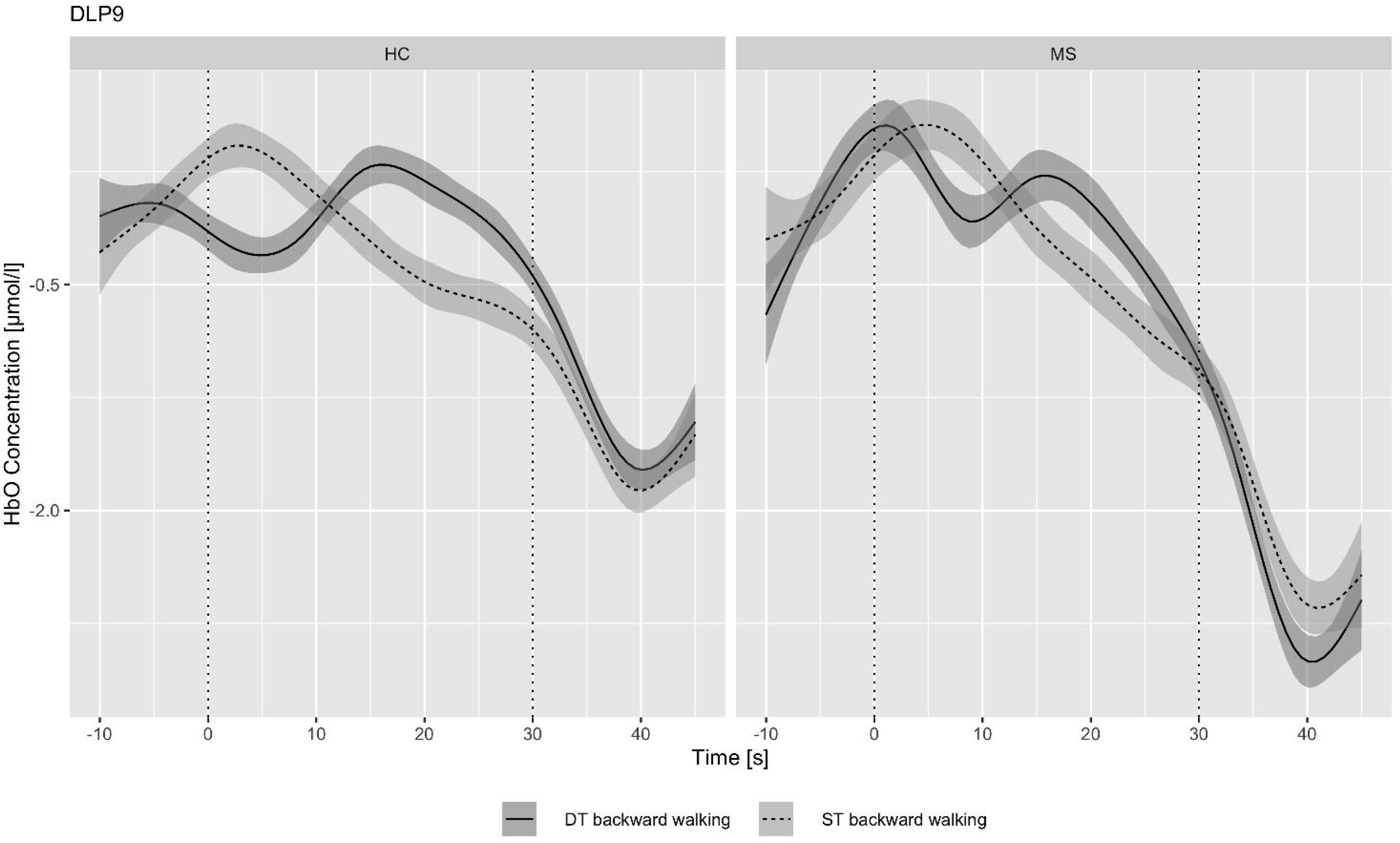
HbO concentration in the DLPFC during ST backward walking and DT backward walking in pwMS (MS) and healthy controls (HC)

**Fig. 3b.**
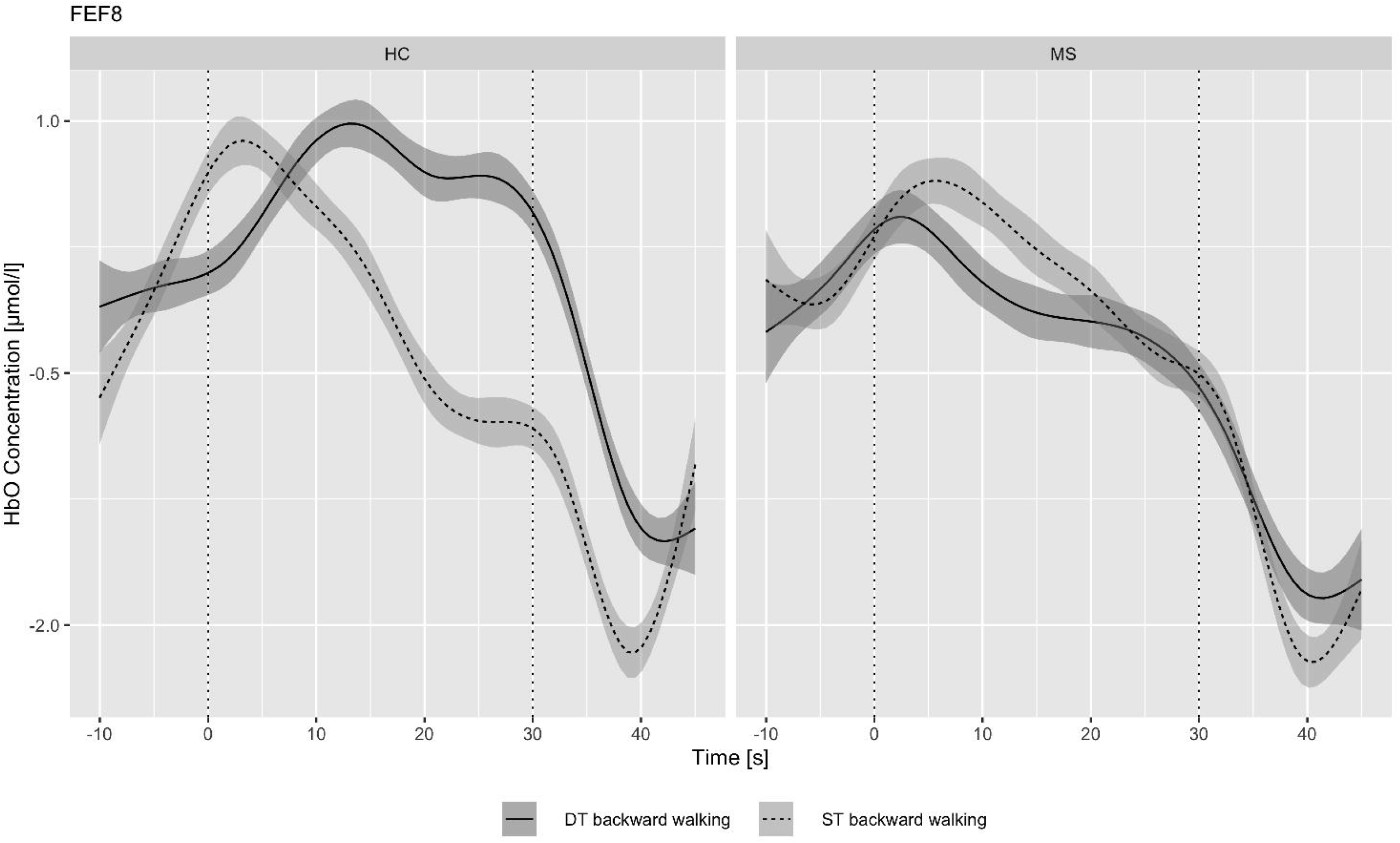
HbO concentration in the FEF during ST backward and DT backward walking in pwMS (MS) and healthy controls (HC)

**Fig. 3c.**
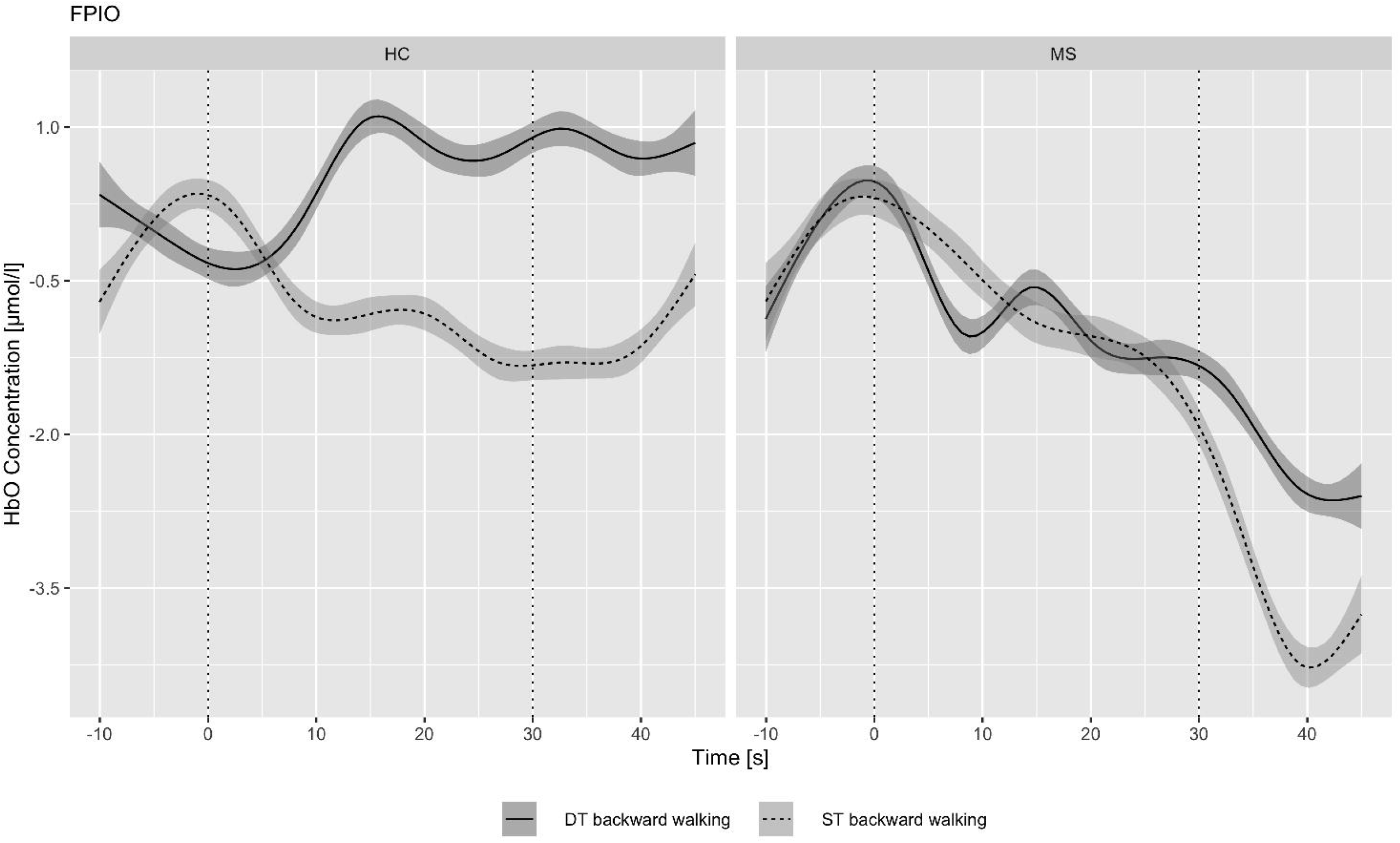
HbO concentration in the FPC during ST backward walking and DT backward walking in pwMS (MS) and healthy controls (HC)

## Discussion

The primary aim of the present study was to investigate the differences in PFC hemodynamics during ST and DT forward as well as backward walking in pwMS. To the best of our knowledge, this study is the first to investigate these differences, specifically, in respect of backward walking. Regarding our first hypothesis, the relative HbO concentration was higher during DT forward walking compared with ST forward walking in the FPC and FEF subareas for both groups but more pronounced in the healthy controls. As for our second hypothesis, the relative HbO concentration was higher during ST backward walking compared with ST forward walking in pwMS and healthy controls, specifically, during the initial part of the walking trial. Non-significant differences were observed between DT backward and DT forward in both groups.

Only a few previous studies have examined PFC activity via fNIRS while walking in pwMS,^20,24,40,50^ with only two studies demonstrating comparable results with our study, since they include ST and DT walking conditions.^20,24^ Chaparro et al examined PFC activation during treadmill walking (with/without weight bearing support) under ST and cognitive-motor DT conditions in a small group of pwMS (n = 10; EDSS = 3.7) and healthy controls (n = 12). Their main finding was a significantly higher HbO concentration during the DT walking condition (without weight bearing support) in pwMS compared with healthy controls.^22^ The same research group investigated the HbO concentration in the PFC during over ground ST and DT walking in a small sample of pwMS (n = 8) and older adults (n = 8).^24^ The HbO concentration in the PFC under ST and DT conditions were higher in pwMS compared to healthy adults. Nonetheless, the difference in the HbO concentration between ST to DT walking was less profound in pwMS classified as mildly disabled (EDSS < 4) compared with patients classified as moderately disabled.

These results are similar to our findings to the extent that the addition of a cognitive task to forward walking causes an increase in PFC activity. Nevertheless, our findings indicate that the increase in PFC activity during motor-cognitive DT is not much different between mildly disabled pwMS and healthy adults. The increased PFC activation is probably related to its role in key cognitive functions in pwMS. The circuit comprising the PFC, along with the caudate, globus pallidus and thalamus, plays a putative role in regulating attention and executive functions in pwMS.^51^ Therefore, the higher HbO concentration demonstrated during DT walking compared with ST walking, suggests an effective adaptive mechanism needed to deal with this relatively complex task in pwMS, particularly those patients with cognitive dysfunctions.^12-14^

An observation from our study is that the relative HbO concentration in the PFC often exhibited negative values during ST forward walking indicating a higher PFC activity during the baseline. This finding is consistent with prior literature showing reduced or very small increases in PFC activity during ST walking relative to baseline.^52-54^ This may be explained by heightened levels of attention and planning during task preparation, which then dissipates as automatic control processes at lower levels of the neuraxis take over during task performance.^52-53^ Another possible factor is that the HbO delivery to the PFC is reduced as blood is diverted to other regions (e.g., motor and visual cortex) that are important for walking.^55^

To date, there are limited and controversial data relating to brain activity during backward walking in pwMS, with the majority of studies focusing on related motor areas (e.g. supplementary motor area).^56-58^ With respect to PFC activity during backward walking, there are only studies on other populations such as healthy adults. Berchicci et al examined PFC activity (via EEG) during self-paced forward and backward stepping in a small group of young healthy adults (n = 11). They demonstrated that the activity in the PFC was elevated compared to baseline during forward and backward stepping tasks, but, was especially, enhanced for the latter.^59^ Recently, Takami et al. investigated changes in the PFC hemodynamics (via fNIRS) during ST forward and backward (treadmill) walking with speed misperception generated by virtual reality (VR). The authors found that during backward walking (after VR viewing), there was an increase in the PFC hemodynamics (mainly, the right region) compared with forward walking.^60^

Although the measurement systems and test settings diverge from our study (EEG vs. fNIRS and treadmill vs. overground walking), the results are similar indicating additional PFC activity during ST backward walking compared with ST forward walking. A possible interpretation might be the different cognitive load due to the complexity of the task. Forward walking is highly automatized in contrast to backward walking.^61^ Thus, backward walking is more cognitively demanding and requires more attentional resources as well as executive control.^62^ This has already been shown for other populations such as elderly and people with neurodegenerative diseases.^63,64^

Worth noting, the difference between backward walking and forward walking in our study was mainly reflected in a higher activation of the FPC and the FEF. The FPC plays a central role in higher cognitive functions such as planning, problem solving, and reasoning. The FEF is involved in motion information processing. The difference between forward and backward walking was less consistent in the DLPFC. Accordingly, future studies related to backward walking should further investigate the differences between the subareas of the PFC.

One general limitation of this study was that all included pwMS suffered from a relapsing-remitting clinical disease course, hence, the present results may not be specifically generalized to pwMS afflicted with a progressive disease type. Another technical limitation was that only one cap size (58 cm circumference) was used for all subjects. This may have led to inaccuracies in the measurement results.

Future studies should consider analyzing also other related brain areas such as the premotor cortex, supplementary motor area or the primary motor cortex to provide a deeper understanding of the underlying mechanisms. Unfortunately, this option was not possible with the fNIRS device applied in this study. Furthermore, in addition to the Serial-7 Subtraction Test used in this study, cognitive tests that reflect e.g. the executive functions (working memory, inhibitory control and cognitive flexibility^65^ should be examined in more detail, especially in relation to the PFC activation during motor ST and motor-cognitive DT.

## Conclusions

The current study presents innovative data on PFC activity during ST and motor-cognitive DT forward and backward walking in pwMS. According to our data these walking conditions impact the hemodynamics at the PFC, although, the difference between pwMS and healthy adults requires further clarification. Our findings should encourage future randomized clinical trials to examine the impact of an intervention program based on DT forward and backward walking on PFC activity along with cognitive function in the pwMS.

## Data Availability

All data produced in the present study are available upon reasonable request to the authors

## Declarations

### Ethics approval and consent to participate

Approval was obtained from the Sheba Medical Center Independent Ethics Committee prior to the commencement of the study (Ethics Ref: 5596-08/141210). All participants provided written informed consent.

### Author contributions

**Yana Kupchenko:** Data curation, visualization, formal analysis, writing review and editing (equal).

**Sapir Dreyer-Alster:** Data curation, visualization, formal analysis, writing review and editing (equal).

**Kim-Charline Broscheid:** Conceptualization, resources, supervision, writing review and editing (equal).

**Alon Kalron:** Conceptualization, formal analysis, methodology, supervision, writing original draft.

## Acknowledgements

The authors thank Mr. Lior Frid for his assistance in data collection.

## Availability of data and Materials

Upon request from corresponding author

## Legends to figures

For all figures the lines (complete and dotted) represent the mean value of the walking trial condition. The shaded area represents the standard deviation.

